# Current challenges of severe acute respiratory syndrome coronavirus 2 (SARS-CoV-2) seroprevalence studies among blood donors: A scoping review

**DOI:** 10.1101/2021.05.13.21257177

**Authors:** Sahar Saeed, Samra Uzicanin, Antoine Lewin, Ryanne Lieshout-Krikke, Helen Faddy, Christian Erikstrup, Carla Osiowy, Clive Seed, Whitney Steele, Katy Davidson, Brian Custer, Sheila O’Brien, On behalf of the Surveillance Risk Assessment and Policy (SRAP) sub-group of the Transfusion Transmitted Infectious Diseases Working Party of the International Society of Blood Transfusion

## Abstract

**Background:** Blood donors are increasingly being recognized as an informative resource for surveillance. We aimed to review and characterize SARS-CoV-2 seroprevalence studies conducted using blood donors to investigate methodology and provide guidance for future research.

**Methods:** We conducted a scoping review of peer-reviewed and preprint publications between January 2020 to January 2021. Two reviewers used standardized forms to extract seroprevalence estimates and data on methodology pertaining to population sampling, periodicity, assay characteristics and antibody kinetics. National data on cumulative incidence and social distancing policies were extracted from publicly available sources and summarized.

**Results:** Thirty-three studies representing 1,323,307 blood donations from 20 countries worldwide were included (sample size per study ranged from 22 to 953,926 donations). Seroprevalence rates ranged from 0% to 76% (after adjusting for waning antibodies). Overall, less than 1 in 5 studies reported standardized seroprevalence rates to reflect the demographics of the general population. Stratification by age and sex were most common (64% of studies), followed by region (48%). 52% of studies reported seroprevalence at a single time point. Overall, 27 unique assay combinations were identified, 55% of studies used a single assay and only 39% adjusted seroprevalence rates for imperfect test characteristics. Among the eight nationally representative studies case detection was most underrepresented in Kenya (1:1264).

**Conclusion:** As of December 11, 2020, 79% of studies reported seroprevalence rates <10%; thresholds far from reaching herd immunity. In addition to differences in community transmission and diverse public health policies, study designs and methodology were likely contributing factors to seroprevalence heterogeneity.

## Introduction

As health authorities contend with the unrelenting coronavirus disease 2019 (COVID-19) pandemic, resources continue to be invested in tracking population-level exposure to severe acute respiratory syndrome coronavirus 2 (SARS-CoV-2). Case detection can be used to monitor infection rates but may underestimate prevalence by limited testing capacity; the restricted time period SARS-CoV-2 is detectable by diagnostic tests; and a significant proportion of mildly and asymptomatic cases that do not seek testing. In contrast, serological tests that identify SARS-CoV-2 specific antibodies are commonly used for surveillance studies, overcoming the limitations of relying on case detection alone.

Given the unprecedented urgency to evaluate the burden of COVID-19, SARS-CoV-2 seroprevalence studies were mobilized quickly. While in theory, random sampling from the general population (e.g., population-based seroprevalence studies) should yield the most generalizable results, this approach is both time-consuming and expensive. Additionally, time-varying response rate may lead to a complex selection bias. In contrast, populations of blood donor have increasingly been recognized as an informative and cost-effective strategy to monitor epidemics [1]. Blood services have the operational capacity to sample and test large proportions of the healthy population for surveillance purposes [2-4]. The Surveillance Risk Assessment and Policy (SRAP) sub-group of the Transfusion Transmitted Infectious Diseases Working Party (TTIDW) of the International Society of Blood Transfusion (ISBT) recently published 73% (32/48) of blood operators surveyed worldwide were undertaking or planning to conduct seroprevalence studies to inform public health [5].

Methodological challenges have emerged unique to this pandemic. The validity, interpretability, and ability to pool seroprevalence studies are limited by study designs, sampling strategies, study timing, the variability in assay characteristics and antibody kinetics. Seroprevalence studies, including those among blood donors, are compiled by on-line dashboards such as SeroTracker (https://serotracker.com) [6], and editorials and perspective articles have been published on limitations of seroprevalence studies in general [1, 7, 8], but to our knowledge there has not been an attempt to systematically bridge these two elements together to provide epidemiological guidance for future research. In this scoping review, we summarized studies specifically among blood donors to characterize SARS-CoV-2 seroprevalence studies, evaluate how well subpopulations and geographic areas have been represented, and determine the diversity of methodology used to address limitations associated with these studies. Given the scope of the research aims, expected heterogeneity and the urgency to inform methodology and reporting of future seroprevalence reports we conducted this synthesis.

## Methods

This scoping review was conducted and supplemented with publicly available data on cumulative COVID-19 cases and social distancing policies nationally. Additionally, members of the ISBT TTIDWP (representing blood collectors from Canada, United States, Denmark, The Netherlands, and Australia) provided expert opinions. Findings were reported according to the Preferred Reporting Items for Systematic Reviews and Meta-Analysis extension for Scoping Reviews (PRISMA-ScR) [9]. Given the time frame a review protocol was not registered.

Two reviewers (SS and SU) independently searched articles using the search engines PubMed and medRxiv for SARS-CoV-2 seroprevalence studies in English among blood donor populations from January 1, 2020 to January 19, 2021. Search terms were [SARS-CoV-2] AND/OR [COVID-19] AND [seroprevalence] AND [donor] OR [blood donor]. A seroprevalence survey was defined as the serological testing of residual blood from blood donations over a restricted time period, to estimate the prevalence of SARS-CoV-2 antibodies in a specified population. Therefore, only studies that reported the sample size, sampling dates and prevalence estimates (or the number of reactive samples) were included in this review. They excluded studies that used residual blood from convalescent plasma donors or as negative controls to evaluate assay performance. Seroprevalence estimates from the grey literature were not included in this review since methods are not routinely reported.

Articles were screened on titles and abstracts by the same two authors. The full-text assessment and data extraction were performed by one reviewer per article and subsequently checked for accuracy by a second reviewer. Consensus was reached between the authors when discrepancies arose. Data were entered into Microsoft Excel using a standardized form, which included: the full reference, region, data of sample collection, data necessary to calculate unadjusted SARS-CoV-2 seroprevalence (the number of samples tested and reactivity). When available the adjusted seroprevalence estimate(s) and 95% confidence intervals (CI) or a range of estimates were extracted. The authors *a priori* identified specific challenges of seroprevalence studies (graphically represented as **Figure 1**):

1. Population sampling: What was the scope of the study, national or regional? Were blood donor populations characterized? Were seroprevalence rates stratified by age, sex, socioeconomic status or by specific regions? Was the seroprevalence estimate standardized to population level characteristics?
2. Dynamic epidemic: What was the type of study design (single or serial cross sectional) to evaluate temporal trends?
3. Assay characteristics: Was the assay reported? Was the assay commercial or an in-house assay? Were the seroprevalence estimates adjusted for imperfect assay characteristics? And how?
4. Antibody kinetics: Were estimates adjusted for waning antibody titers?

**Figure 1:**
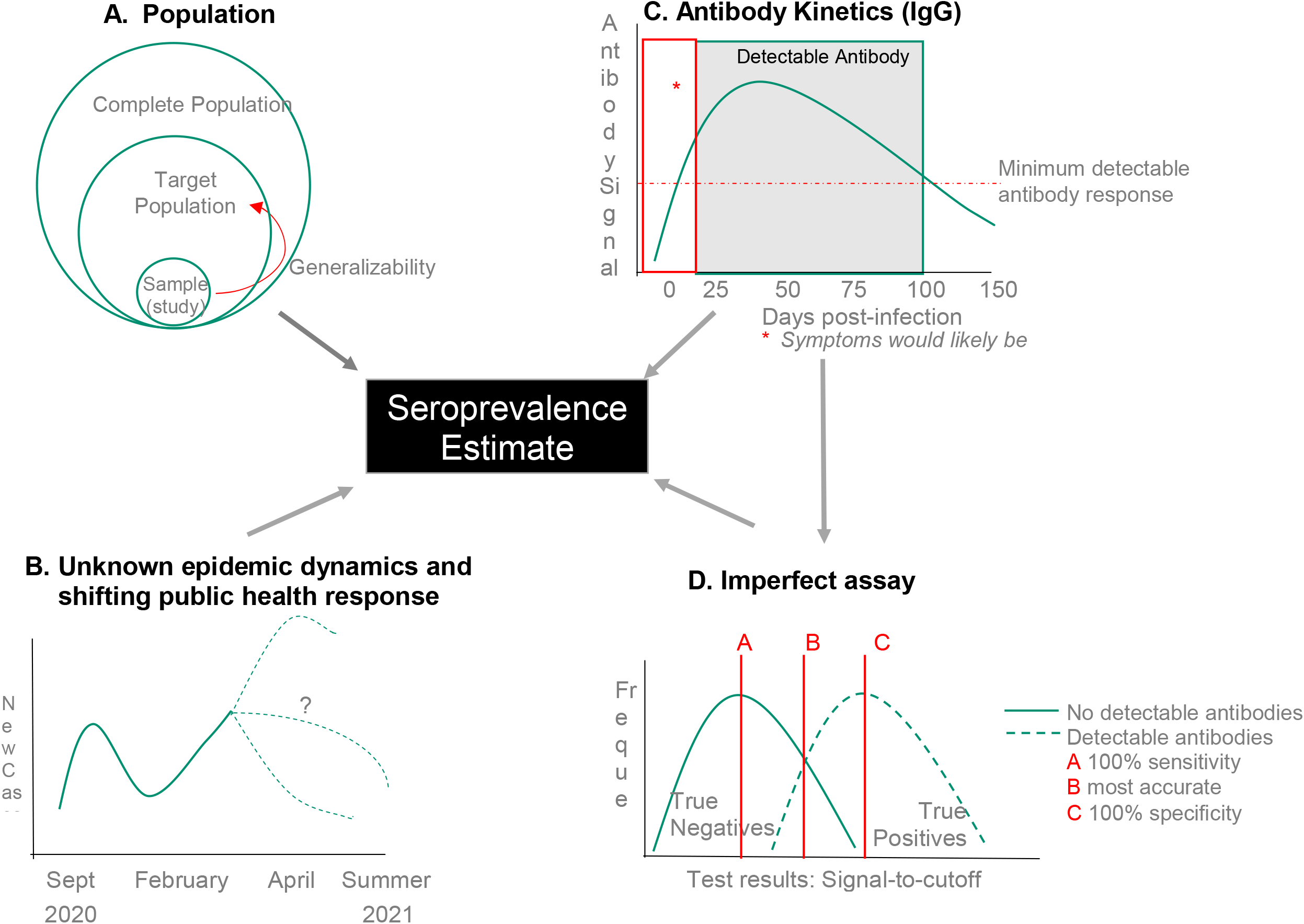
Overlapping determinants of estimating population-level seroprevalence. **A. Population**– The WHO endorses seroprevalence studies of blood donors since they are a convenient sample of healthy adult population. But care should be taken when generalizing the results beyond the target population. Additionally, since infection rates are likely differential by regions, socioeconomic status, age and racialized populations minorities within a country, grouping prevalence rates into a single summary may miss significant differences. There is also a potential for selection bias as donors are a self-selected group of people. **B. Shifting Public Policy**– Seroprevalence can be influenced by changing population-level trends, defined as surges of new cases followed by a downturn (epidemic waves) **C. Antibody Kinetics**-From the time of infection, on average, it takes 10-28 days to develop specific immunoglobulin G (IgG) antibodies to SARS-CoV-2. And by approximately 100 days, the level of these antibodies detectable in the blood begins to decrease (wane)^6^. If sampling occurs outside this window of detection, people both early and later in their infection will be missed. **D. Accuracy of Tests**– Test performance is measured by the proportion of people who are accurately identified as having antibodies “sensitivity” and not having antibodies “specificity”, at a given threshold (signal-to-cutoff ratio).

We extracted cumulative case counts from the World Health Organization Coronavirus Disease (COVID-19 Dashboard (https://covid19.who.int/). Among the nationally representative studies we estimate the ratio of reported to expected infections. Since seroprevalence data reflects infections that occurred prior to the date of measurement, case detection was extracted two-weeks from the end of the donation collection/study period for each study [10].

Public health policies were summarized using the Government Stringency Index [11]. This composite measure was based on nine response indicators including school closures, workplace closures, and travel bans, rescaled to a value from 0 to 100 (100 = strictest). If policies varied at the subnational level, the index is shown as the response level of the strictest sub-region. We summarized policy stringency as the average of the daily index two-weeks before the beginning and end of the study period.

## Results

From 157 articles (32 peer-reviewed and 125 pre-prints), 33 studies (22 peer-reviewed and 11 preprints) were included in this review (**Figure 2**). The 33 studies represented 20 countries, the majority from Europe (n=8) followed by North America (n=4), Asia (n=4), Africa (n=2), South America (n=1) and Australia (n=1) (**Figure 3**). Seroprevalence studies from low- and middle-income countries were limited.

**Figure 2:**
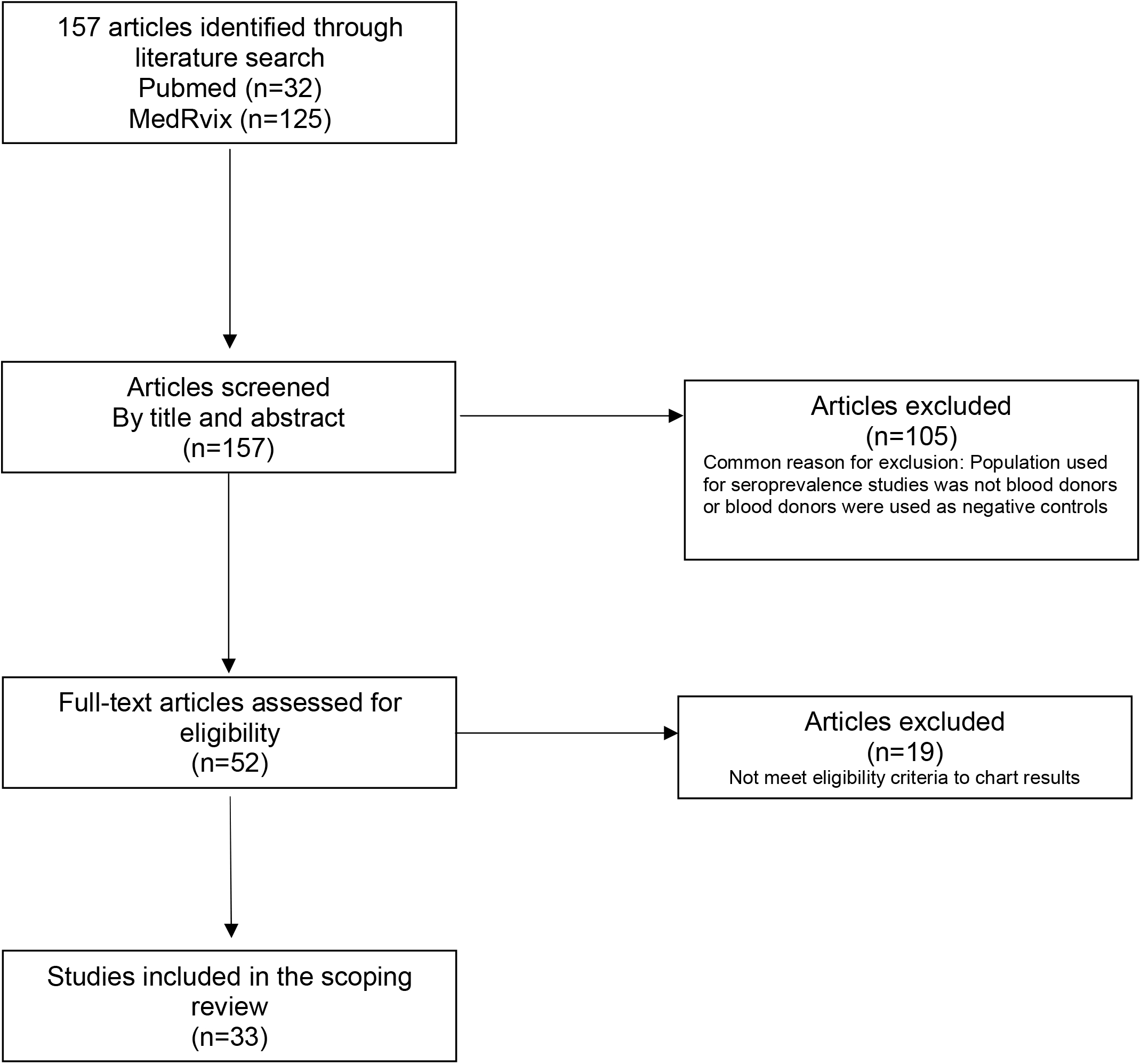
Inclusion of studies in review.

**Figure 3:**
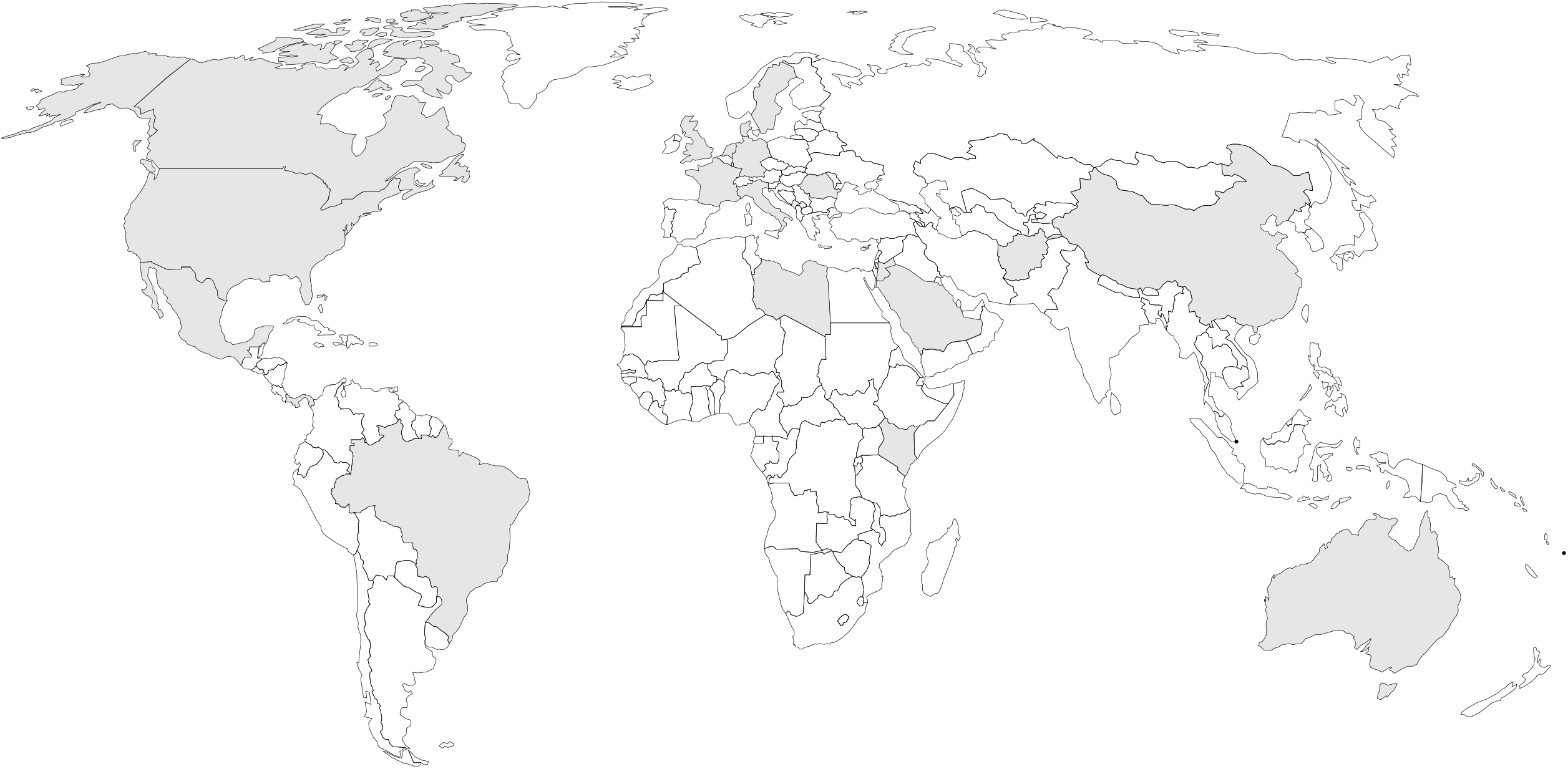
Worldwide coverage of seroprevalence studies among blood donor populations. Shaded countries are represented in this review

**Table 1** characterizes the data extracted from each of the 33 studies included in this review [12-44]. Of the published studies the vast majority (91%; 20/22) were published in clinical or public health journals, two were specifically published in transfusion medicine journals. The median sample size was 1,996 donations but ranged from as many as 953,926 in the USA [28]to as few as 22 in Libya [41]. The time frames for studies ranged from beginning January 1, 2020 to ending December 11, 2020. Nine (27%) studies began between January and February 29, 2020; one study began in September 2020. **Figure 4** illustrates seroprevalence rates ranging from as low as 0% in Saudi Arabia to as high as 34-37% in Pakistan. One study from Brazil reported unadjusted seroprevalence rates of 25.8%% but after adjusting for waning antibodies seroprevalence rates were as high as 76% [43].

**Table 1.**
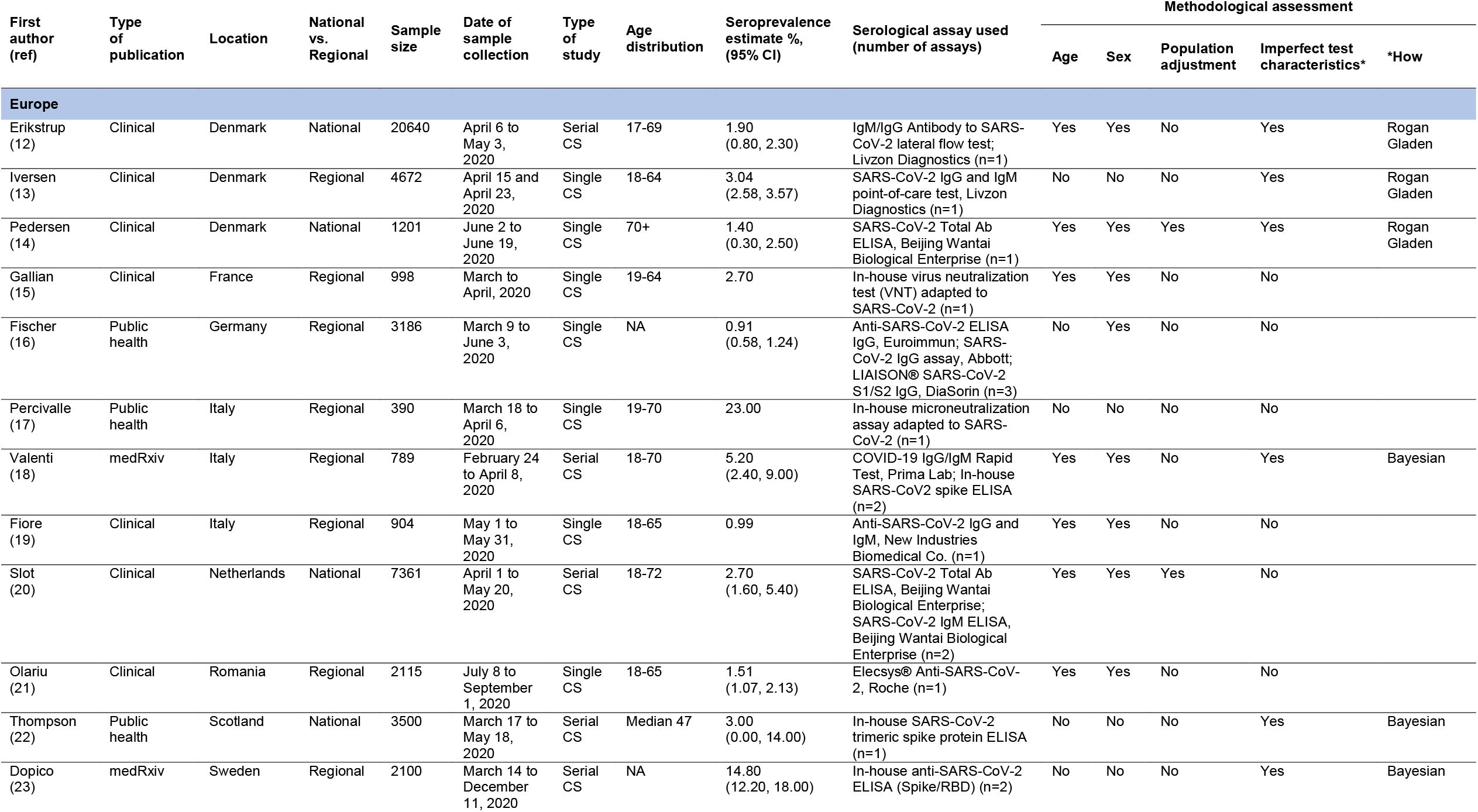
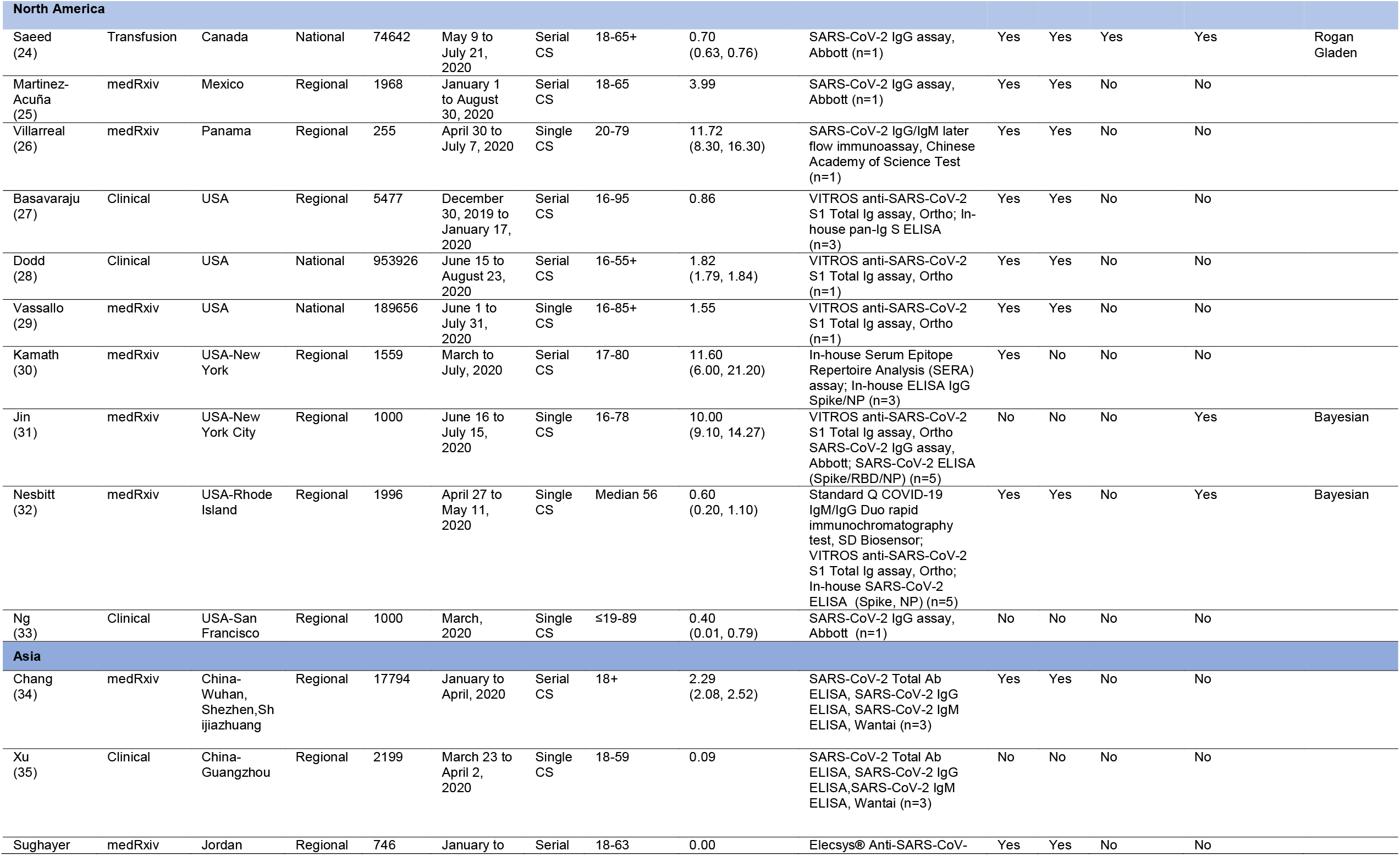
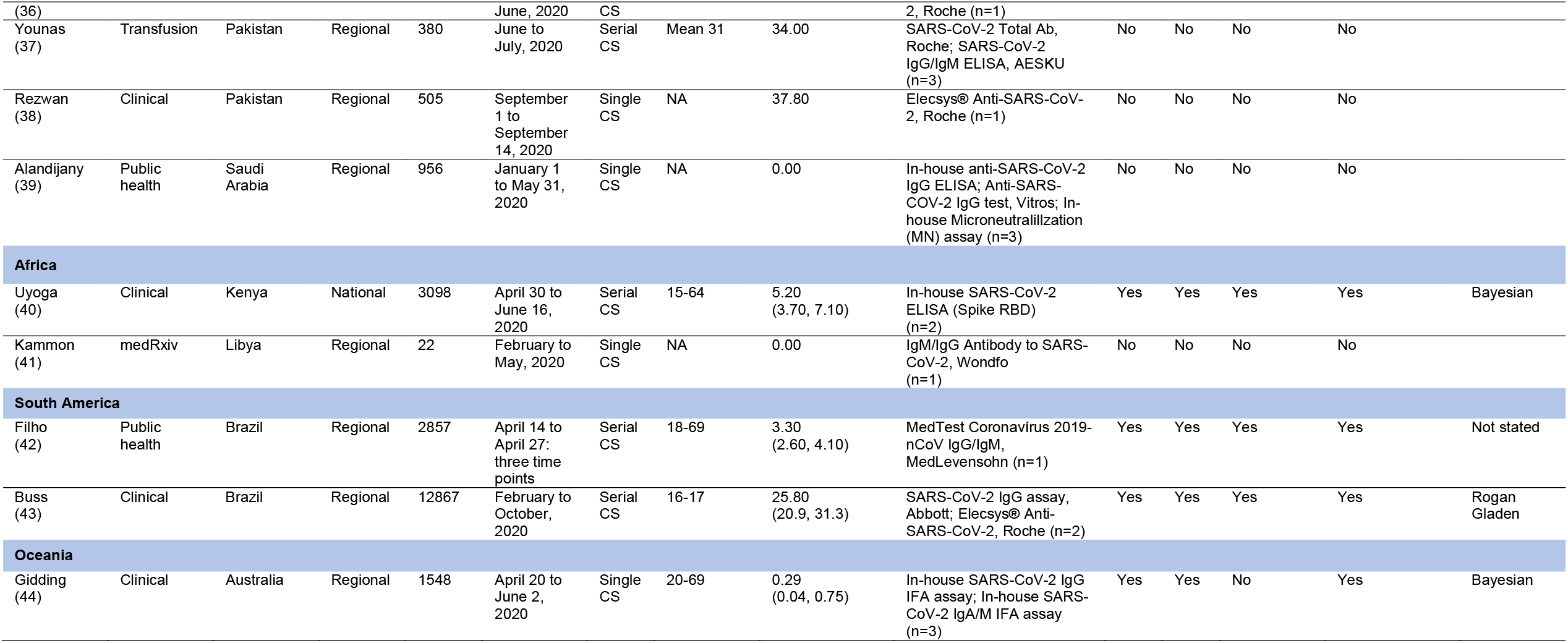
Characteristics of studies included in this review (n=33)

**Figure 4:**
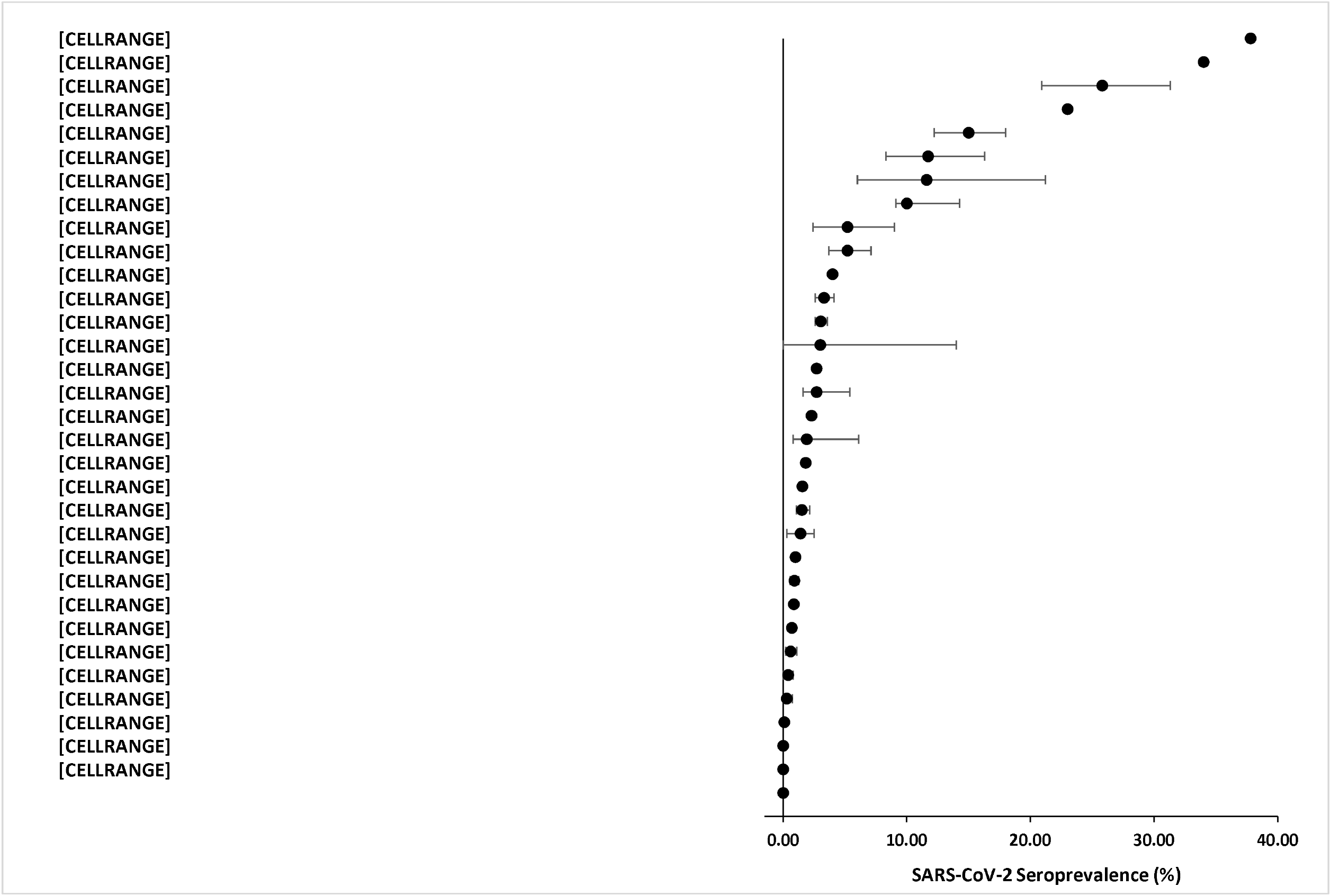
Forest plot of seroprevalence estimates. Seroprevalence estimates from each study is ranked from highest to lowest. Regional estimates within countries are specified in brackets. Bars around the estimates represent 95% confidence intervals (when available) or estimate ranges (Scotland). ^**^Seroprevalence unadjusted for waning antibodies (76% if adjusted)

There was significant heterogeneity by methodological factors that influence seroprevalence estimates (**Table 1, summarized by Figure 5**). Characterization of the blood donor populations varied; five studies did not report the age of donors sampled [16, 23, 38, 39, 41]. The scope of studies varied, the majority (76%; 25/33) provided regional estimates within countries. Approximately half of the studies (52%; 17/33) provided a single seroprevalence estimate [13-17, 19, 21, 26, 29, 31-33, 35, 38, 39, 41, 44]. Stratification by age and sex were most common (64%; 21/33), followed by region (48%; 16/33). While age stratification was common, there was no standardized age grouping. Very few studies stratified by a broad definition of socioeconomic status (15%; 5/33) (definition of SES varied: i.e. income, occupation, or neighborhood-level social and material deprivation) [24, 25, 29, 34, 42]. Overall, less than 1 in 5 studies adjusted seroprevalence rates to reflect the demographics of the general population. There were almost as many unique assay combinations (n=27) as studies included in the review. A single assay was used most often (55%; 18/33), 15 studies used two or more assays to determine seroprevalence estimates. Of the 18 studies used a single assay [n=15 different assays]; 15 studies used two or more assays [n=12 different assay combinations]). At least one in-house assay was used by 33% of studies. Overall, 39% of studies adjusted seroprevalence estimates by imperfect test characteristics; (38%; 5/13) of the studies used the Rogan-Gladen [12-14, 24, 43] equation to adjust for imperfect sensitivity and specificity and (54%; 7/13) used Bayesian methods [18, 22, 23, 31, 32, 40, 44] and one study did not state the method for adjustment [42]. Only two studies (Sweden and Brazil), adjusted their seroprevalence estimates for waning antibodies [23, 43].

**Figure 5:**
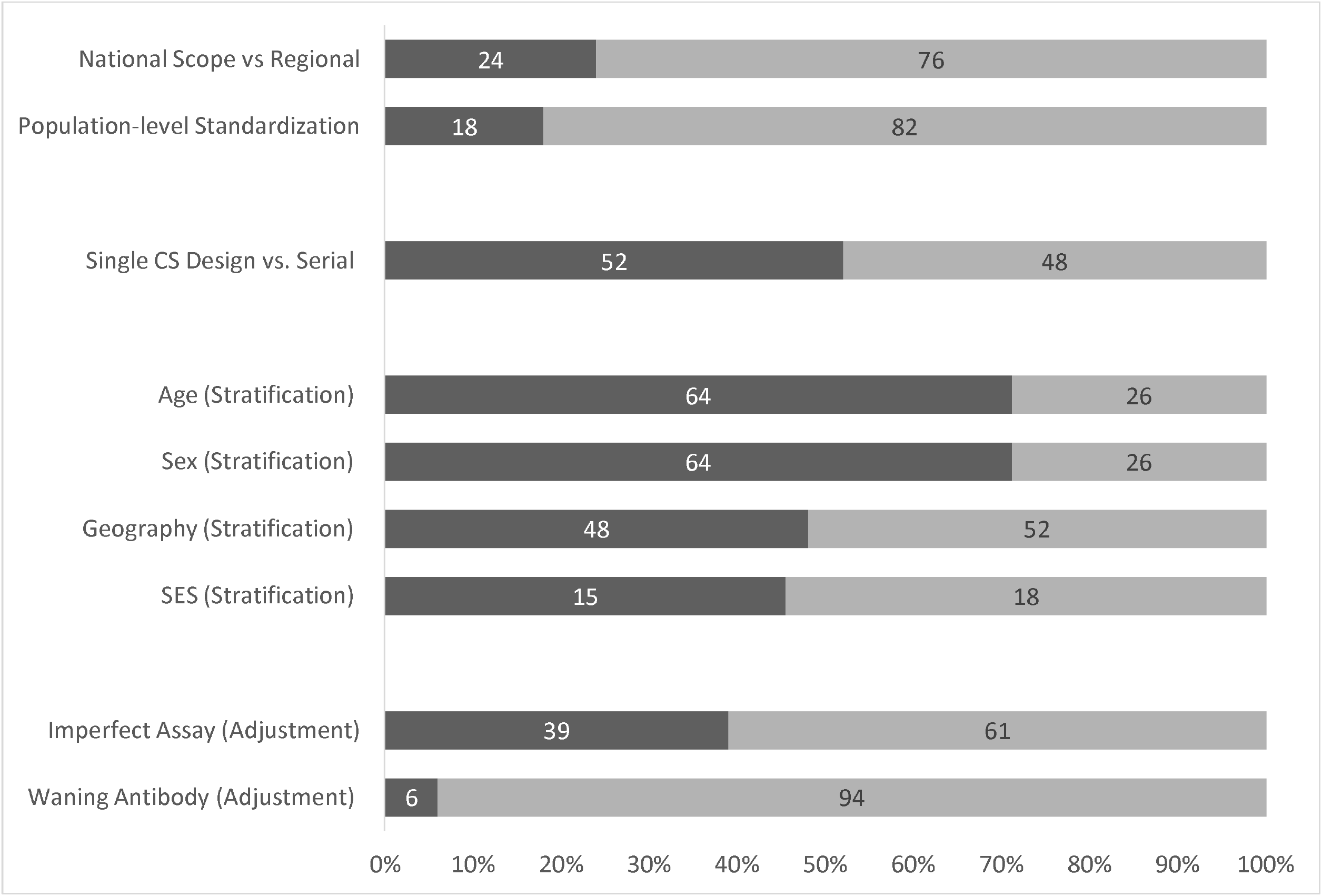
Summary of analytical considerations. CS-Cross-sectional; SES-socioeconomic status

Based on the ratio of reported to expected cases, we found case detection was most underreported in Kenya (0.001=1:1264) (**Table 2**). Among the eight nationally representative studies, the policy stringency index during the study period ranged from 59.37 to 88.89 (**Table 2**). We found no linear association between seroprevalence rates and policy stringency.

**Table 2.** Ratio of reported infections to seroprevalence-derived number of infections among nationally representative study’s (n=8)

| Country (ref) | Population in 2020 | Average policy stringency | Seroprevalence estimate %, (95% CI) | Infections 14 days prior to end of the study (cumulative cases) | Seroprevalence-derived expected number of infections | Seroprevalence-derived expected number of infections range | Ratio of reported infections to seroprevalence-derived number of infections |
| --- | --- | --- | --- | --- | --- | --- | --- |
| Denmark (12) | 5,792,202 | 71.45 | 1.90 (0.80, 2.30) | 7,384 | 110,052 | 46,338, 133,221 | 0.067 |
| Denmark (14) | 5,792,202 | 59.37 | 1.40 (0.30, 2.50) | 11,771 | 81,091 | 17,377, 144,805 | 0.145 |
| Netherlands (20) | 17,134,872 | 77.65 | 2.70 (1.60, 5.40) | 40,841 | 462,642 | 274,158, 925,283 | 0.088 |
| Scotland (22) | 5,460,000 | 60.65 | 3.00 (0.00, 14.00) * | 12,226 | 163,800 | 0, 764,400 | 0.075 |
| Canada (24) | 29,247,654** | 70.82 | 0.70 (0.63, 0.76) | 50,169 | 204,734 | 184,260, 222,282 | 0.245 |
| USA (28) | 331,002,651 | 69.11 | 1.82 (1.79, 1.84) | 4,999,815 | 6,024,248 | 5,924,947, 6,090,449 | 0.830 |
| USA (29) | 331,002,651 | 70.71 | 1.55 | 3,405,494 | 5,130,541 | NA | 0.664 |
| Kenya (40) | 53,771,296 | 88.89 | 5.20 (3.70, 7.10) | 2,216 | 2,796,107 | 1,989,538, 3,817,762 | 0.001 |

\*Not a 95% CI instead a range of seroprevalence rates by geographical regions
\*\*Excluding the province of Quebec since not represented in the study

## Discussion

In this scoping review we summarized results from 33 SARS-CoV-2 seroprevalence studies among blood donors, representing 1,323,307 donations worldwide. As of December 11, 2020, 79% of studies reported seroprevalence rates <10%; thresholds far from reaching herd immunity. In addition to variations in community transmission and the diverse public health response to the COVID-19 pandemic, we found study designs and methodology were contributing factors to seroprevalence heterogeneity. As health officials turn to seroprevalence studies to bridge the gap left by diagnostic testing, we provide epidemiological guidance to improve seroprevalence reporting specifically among blood donor populations.

Blood donors are a selected population of a target population (**Figure 1**). To evaluate the generalizability of seroprevalence studies, demographic characteristics of both the study and target populations are necessary. We found, only one in five studies adjusted their donor population to reflect the demographics of the general population [45-47]; a straightforward adjustment that can be accommodated by most statistical software. Furthermore, it is important to note there can be significant variations of an individual’s willingness to donate blood, recruitment strategies and eligibility criteria across blood operators. For example, according to the WHO, donating blood is more common in high-income compared to lower income countries [48]. Additionally, many lower- and middle-income countries rely on family replacement donors to donate on behalf of patients rather than altruistic volunteers. In 2016 it was estimated that nearly 30% of blood donations worldwide, occurred in Europe, which only accounts for 10% of the world’s population. A larger proportion of the population donating blood increases the generalizability of results from studies of blood donors. Indeed, a recent study comparing seroprevalence estimates from European blood donors to household surveys targeting the general population found seroprevalence rates to be very similar [49].

Differential characteristics between study and target populations not only affect the generalizability of the estimates but a selection bias known as the “healthy donor effect” when evaluating predictors of seroprevalence. Briefly donor’s self-selection and eligibility criteria may skew blood donors to be healthier than the general population [50]. Unhealthy donors are likely to self-defer or be deferred at the time of collection, leaving a group of more health-conscious donors. Moreover, it is possible some blood donors may have been incentivized to donate by disclosure of SARS-CoV-2 antibody results. This may distort the sample towards people who believe they were previously infected seeking confirmation. While these factors may have been possible early during the pandemic due to limited testing capacity it is unlikely to pose a significant incentive to donate as immunoassays became more readily available. Addressing selection bias is more complicated and requires additional information that may not be readily available to researchers, such as the data on variables influencing selection and the outcome among non-donors. When these data are available methods such as inverse probability of selection weighting (IPSW) can correct for this bias [51].

There is strong evidence to support differential SARS-CoV-2 infection rates by calendar time, geographic region, age groups and socioeconomic status [52. 53]. Therefore, providing a single seroprevalence estimate may miss significant disparities. While most studies did provide stratified rates by age, there was no standardized age grouping making comparison between regions/countries difficult. Furthermore, very few studies stratified by a broad definition of socioeconomic status and the definition for SES varied considerably. Stratifying seroprevalence estimates by socioeconomic demographics can provide public health officials critical data to implement targeted interventions. Additionally, the granular data can be used to produce more precise estimations of case identification fractions and infection fatality rates by subgroups [54, 55]. Careful consideration should be made when designing studies with sufficient sample sizes to accurately estimate these subgroup analyses. Furthermore, we recommend seroprevalence studies to state their target population and the cumulative incidence reported by case detection to provide readers insight on the magnitude of testing capacity.

The accuracy of the assay, antibody kinetics and population-level epidemic changes are overlapping challenges that affect the validity of seroprevalence estimate (**Figure 1**). No assay is 100% sensitive and specific yet only a third of the studies we reviewed adjusted there seroprevalence estimates for this imperfection. When the sensitivity and specificity are known, seroprevalence estimates can be adjusted using either the Rogen-Gladen equation or using Bayesian methods [56, 57]. Orthogonal testing may also increase accuracy of estimates when multiple assays are available. We found the majority of the studies used a single assay to identify seropositivity. In the absence of a gold standard multiple methods can be applied to correct for this measurement error ranging from deterministic predefined rules to more advanced latent class models [58, 59].

Further consideration when comparing and interpreting seroprevalence studies using different assays include different antibody isotypes and that target various SARS-CoV-2 proteins. For example, the type of antibody isotypes (IgA, IgM or IgG) reflect various stages of an infection. IgG is detectable 10-21 days post infection and remains detectable longer than IgA and IgM. Even though SARS-CoV-2 is likely not transfusion transmission [60, 61], it was common for blood operators to defer donations for 2-4 weeks following resolution of symptoms (after confirmed diagnosis of COVID-19) or were exposed to SARS-CoV-2 through travel or contact [62]. Given this deferral period, the early period of undetectable IgG levels is less likely to be an issue among blood donors, although some early-stage asymptomatic donors may not be detected. Assays vary by detecting specific SARS-CoV-2 proteins (surface or spike glycoprotein (S), spike protein receptor binding domain (RBD), and nucleocapsid (N)) and currently there is no consensus on which antigen is most frequently expressed and for how long. While sensitivity and specificity are independent test characteristics, underlying disease prevalence affects the positive and negative predictive values of assays. In this review we found the majority of studies included were low prevalence settings (<10%) meaning minor fluctuations from perfect specificity would cause inflation of false positive results affecting seroprevalence estimates.

Typically, seroprevalence studies are cross-sectional and occur either at a single time point or serially. Based on consultation of ISBT members conducting ongoing seroprevalence surveys there was no consensus on how often seroprevalence studies should be conducted. However, given the dynamic nature of the pandemic multiple estimates over time would provide more informative results for public health authorities. Single cross-sectional studies are appropriate for chronic infections when the antibodies remain present for extended periods of time. However, there is mounting evidence that by approximately 100 days post infection, anti-SARS-CoV-2 IgG antibodies begin to wane and can fall below levels of detectability [63, 64]. Therefore, depending on the timing of the study, seroprevalence estimates will include a varying proportion of “recent” infections which will mirror epidemiology curves by case detection and “past” infections which may or may not be detectable given a specific threshold. Blood donor seroprevalence studies are used as a proxy of the general population who may not have known they were infected with SARS-CoV-2. Without a known date of infection waning antibody signals makes it difficult to quantify what proportion had previous infections that are no longer detectable. While the studies that focused on the first wave of the pandemic are likely to capture most infections (past and recent), it will become more difficult for future studies to capture cumulative seroprevalence without serial designs adjusting for waning antibodies. Various methods are being explored to adjust for waning antibody signals including increasing sensitivity by lowering sample to cut-off ratio thresholds and by using population-level incidence and mortality data to model approximate infection dates [64, 65]. Given the goal of seroprevalence studies is to determine population-level immunity, more research is needed to identify the correlates of immunity through evaluating waning antibody signals, antibody titers vs. neutralizing capacity and the role of cell-mediated immunity [66]. This will require pairing modeling approaches with confirmatory neutralization assays or a surrogate immunoassay, once confirmed as a correlate [67]. Additionally, longitudinal studies will be required to evaluate the rate of antibody and signal decline. One advantage of conducting seroprevalence studies within blood donor populations is donors tend to donate multiple times a year therefore creating a pseudo-longitudinal cohort that can be used to monitor antibody kinetics.

This review has limitations. It is possible seroprevalence estimates from blood donors were conducted but have yet to be published or possibly never intended to be published as research articles instead reported directly to public health authorities. For example, in Denmark, the Netherlands and England weekly seroprevalence reports were generated from blood operators to inform COVID-19 modelling. Therefore, it is possible our sample is biased towards blood operators and countries who had sufficient resources to prepare findings in the form of a manuscript. We attempted to evaluate the association between non-pharmaceutical interventions and seroprevalence, but this introduced several limitations. First, policies in most countries varied at every-level of government and the policy index uses the most stringent level. This can over represent social distancing policies in countries like the United States were there were significant variations in policies at the federal, state, counties, and individual cities. The stringency policy index weighs each policy equally which may not reflect the impact of the individual policies on incidence. Finally, even though we lagged our seroprevalence rates by two weeks, there is still a possibility of reverse causality (countries that had more cases adopted stricter policies). Considering these limitations, we believe the policy stringency index or other descriptions of policy measures can still provide context to the studies, but associations should be made cautiously. Finally, the methodological limitations reviewed in this study are based on a time when vaccines against SARS-CoV-2 were not available. Now with multiple vaccines approved worldwide, widescale vaccination campaigns are underway, seroprevalence studies will remain to be important to track vaccine induced immunity.

Overall, the results from these studies indicate worldwide seroprevalence rates vary considerably but by the end of 2020, the majority of the countries were far from reaching thresholds to achieve herd immunity. Despite all studies being conducted in blood donors, caution should be exercised when comparing seroprevalence estimates given the significant variability in study designs and methodology as we highlight in this article. Public health authorities mobilized resources quickly and new partnerships were accelerated such as those with blood operators. Despite the limitations we highlight, individual studies have been extremely informative in informing public health authorities and blood donors will continue to play a vital role in facilitating seroprevalence studies to assess and monitor the burden of COVID-19.

## Data Availability

List of references included in this scoping review are available

## Notes

### Competing Interest Statement

The authors have declared no competing interest.

### Funding Statement

Canadian Blood Services

